# Respiratory and Cardiovascular Health Effects of E-Cigarette Substitution: Protocol for Two Living Systematic Reviews

**DOI:** 10.1101/2021.03.18.21253876

**Authors:** Renée O’Leary, Maria Ahmed Qureshi, Giusy Rita Maria La Rosa, Robin W. M. Vernooij, Damian Chukwu Odimegwu, Gaetano Bertino, Riccardo Polosa

## Abstract

**Background:** Despite the clear risks of tobacco use, millions of people continue to smoke. Electronic nicotine delivery systems (ENDS), commonly called e-cigarettes, have been proposed as a substitute for those who are unwilling or unable to quit. Current systematic and narrative reviews on the health effects of ENDS use, respiratory and cardiovascular effects in particular, have come to differing conclusions.

**Objective:** The purpose of our two systematic reviews is to critically assess and synthesize the available human studies on the respiratory and cardiovascular health effects of ENDS substitution for people who smoke. The primary goal is to provide clinicians with evidence on the health effects of ENDS substitution to inform their treatment recommendations and plans. The twin goal is to promote the health literacy of ENDS users with facts on the health effects of ENDS. A secondary goal is to develop policy briefs to provide governmental bodies with a preliminary assessment of the potential public health impacts of ENDS.

**Methods:** These two reviews will be living systematic reviews. The systematic reviews will be initiated with a baseline review. Studies will be evaluated with the JBI quality assessment tools and a checklist of biases drawn from the Centre for Evidence Based Medicine’s Catalogue of Bias. A narrative synthesis is planned due to the heterogeneity of data. A search for recently published studies will be conducted every three months, and an updated review published every six months for the duration of the project or possibly longer.

**Results:** The baseline and updated reviews will be published in a peer review journal. The review findings will be reported in a white paper for clinicians, a fact sheet for people who use ENDS, and country-specific policy briefs.

**Conclusions:** The substitution of ENDS for cigarettes is one of the ways to potentially reduce the risks of smoking. Clinicians and their patients need to understand the potential benefits and possible risks of substituting ENDS for cigarettes. Our living systematic reviews seek to highlight the best and most up-to-date evidence in this highly contentious and fast-moving field of research.

## Introduction

### Background

There are 1.3 billion people worldwide who use tobacco, and more than seven million of them die annually from its use [1]. Up to 11.5% of global mortality can be linked to smoking [2]. For respiratory diseases, smoking is the attributable mortality risk factor for 64.21% of lung, tracheal, and bronchus cancers, 63.44% of larynx cancer, 48.47% of chronic obstructive pulmonary disease (COPD), 15.52% of tuberculosis, 11.93% of asthma, and 11.04% of lower respiratory infections [3]. For cardiovascular diseases, smoking was the attributable risk factor for 34.6% of deaths from aortic aneurysm, 26.8% from peripheral artery disease, 18.41% from ischemic heart disease, and 14.2% from stroke [4]. Smoking is one of the primary acquired risk factors for atherosclerotic disease [4].

Despite the clear risks of tobacco use, millions of people continue to smoke. Smoking has pleasurable effects [5], and some smoke for emotional regulation or to self-medicate their symptoms of schizophrenia or Parkinson’s disease [6]. For those who want to quit, the success rate for cessation attempts is low - approximately 7% at six months [7, 8]. Furthermore, presently around 70% of the world’s population has no access to appropriate tobacco cessation services [9]. Quitting smoking is difficult, many have no support to quit, and some people do not wish to quit.

Electronic nicotine delivery systems (ENDS), commonly called e-cigarettes, have been proposed as a substitute for those who are unwilling or unable to quit [10, 11]. A review by the US National Academies of Sciences Engineering and Medicine [12] states *There is substantial evidence that except for nicotine, under typical conditions of use, exposure to potentially toxic substances from e-cigarettes is significantly lower compared with combustible tobacco cigarettes* (p. 198). The acceptability of ENDS among people who smoke is demonstrated by its rapid uptake: in 2018 there were 41 million people using ENDS compared with 7 million users in 2011 [13]. Clinicians want to know the health effects of ENDS use, asking *“are e-cigarettes marginally safer, thus still too risky to substitute for combustible products, or are they substantially safer?*” [14, p. 1002].

### Prior Reviews – Respiratory Effects

Three recent systematic reviews have been published on the respiratory effects of ENDS. Bals, Boyd [15], prepared for the European Respiratory Society, is based on studies published through August 2016, rendering it out of date. Wang, Liu [16] include exclusively in vitro and in vivo studies, not human studies. Goniewicz, Miller [17] analyzes only cross-sectional studies of risk. Although they found a 40% reduction in adverse pulmonary outcomes, cross sectional studies can only demonstrate an association, not causation.

Four narrative reviews on the pulmonary effects of ENDS have been published since 2019; they have been conducted with a combination of in vitro, in vivo, emissions toxicology, and human studies. The reviewers come to diametrically opposed conclusions on their respiratory effects. Gotts, Jordt [18] and Miyashita and Foley [19, conference presentation not peer reviewed] conclude that there is sufficient evidence of respiratory harm from ENDS use. To the contrary, Traboulsi, Cherian [20] present evidence of both beneficial and adverse effects for people who smoke, while Polosa, O’Leary [21, disclosure: RP and RO are co-authors] argue that ENDS substitution results in primarily beneficial health effects.

### Prior Reviews – Cardiovascular Effects

Claims of strong evidence for long-term systematic effects of ENDS on the cardiovascular system have been made, but reviews in this area have also arrived at conflicting conclusions. One systematic review of in vitro, animal, and human studies presents the conclusion that “most studies suggest potential for cardiovascular harm” caused by sympathetic nerve activation, oxidative stress, endothelial dysfunction, and platelet activation [22, abstract]. Another systematic review found no indications of a significant increase or reduction in cardiovascular disease outcomes (stroke, myocardial infarction, and coronary heart disease) among former smokers who transitioned to ENDS based on cross-sectional population studies of ENDS users [17]. Benowitz and Fraiman [23] conclude that ENDS use is likely to incur lower cardiovascular risks than cigarette smoking based on toxicity studies, known mechanisms, and laboratory models. Two current systematic reviews of human studies on the cardiovascular effects of ENDS display more agreement on study evidence, finding lower acute effects and no chronic increases in heart rate and blood pressure for ENDS use compared with smoking [24, 25].

On the other hand, some reviewers claim that evidence is lacking or insufficient. The National Academies of Sciences, Engineering, and Medicine (NASEM) stated that *there is* ***no available evidence*** *whether or not e-cigarette use is associated with clinical cardiovascular outcomes (coronary heart disease, stroke, and peripheral artery disease) and subclinical atherosclerosis (carotid intima-media thickness and coronary artery calcifications)* [12, p. 7 emphasis in original]. D’Amario et al. [26] state that there is a lack of clear or conclusive data on ENDS use and cardiovascular health. MacDonald and Middlekauff [27] conclude their narrative review of human and emissions toxicology studies by saying *the effects of ECs on long-term cardiovascular health are inconclusive, but concerning* (p. 172). Buchanan, Grimmer [28] in their narrative review of pre-clinical and clinical studies even dismiss the available literature: *While the current but still limited literature suggests that e-cigarette use may lead to fewer negative cardiovascular effects than conventional cigarettes, our review supports that there is not sufficient data to conclusively make these resolutions* (p. 47).

An umbrella review [29] (review of reviews) synthesized seven systematic reviews, but did not include the NASEM review. The umbrella review included three reviews conducted in 2016 or prior when substantially fewer studies had been published, and one review that contributed only two case studies from cannabinoid ENDS use. The remaining three systematic reviews have been discussed in the prior paragraphs. The reviewers suggest that ENDS may provide a strategy for harm reduction with the caveat of the need for more studies.

### Research question

Ascertaining the real-life effects of ENDS substitution on respiratory and cardiovascular health is complex. The complexity is due to how ENDS are frequently used in combination with conventional cigarettes, the differences in ENDS products, variations in the nicotine concentration of liquids, and the varying levels of daily exposure. There is an urgent need for a current systematic review of research given the disagreements among prior reviews. Many of the available reviews have limitations arising from their reliance on non-human study data. Furthermore, many reviews show evidence of biased reporting. In addition, findings from recently published studies not covered in these reviews may render their conclusions obsolete. By conducting two living systematic reviews we are aiming to answer the question: “What are the respiratory and cardiovascular health effects resulting from the substitution of ENDS for conventional cigarettes?”

### PICO criteria

#### Population

adults who smoke cigarettes.

#### Intervention

substitution of ENDS for cigarettes.

#### Comparator

participants who continue to smoke, baseline changes in respiratory/cardiovascular tests of study participants who substitute ENDS for smoking (within-subject), or comparisons to documented smoking outcomes.

#### Outcomes respiratory

changes in chronic cough, phlegm, wheeze, dyspnoea, exacerbations of asthma and COPD, or changes in testing including Forced Expiratory Volume (FEV), chest X-rays, and CT scans.

#### Outcomes cardiovascular

measures of cardiovascular function including blood pressure, heart rate, carotid intima-media thickness and coronary artery calcifications, or changes in cardiovascular disease symptoms (clinical observation or self-report).

### Objectives

The purpose of our two systematic reviews is to critically assess and synthesize the available human studies on the respiratory and cardiovascular health effects of ENDS substitution for people who smoke. We will provide an even-handed assessment of the data, considering all health effects both potentially adverse or beneficial. The primary goal is to provide clinicians with evidence on the health effects of ENDS substitution for people who smoke to inform their treatment recommendations and plans. The twin goal is to promote the health literacy of ENDS users with facts on the health effects of ENDS with a white paper drawn from the review. A secondary goal is to develop policy briefs to provide governmental bodies with an assessment of the potential public health impacts of ENDS on their population of people who smoke. These two systematic reviews will be conducted with the living methodology to keep the information up-to-date and complete, as described below.

## Methods

The protocol conforms to the PRISMA-P requirements [30]; the completed checklist is in Supplementary Materials. This protocol has been submitted for registration with PROSPERO on February 23, 2021. Any deviations from this protocol will be reported in the reviews. These reviews are being conducted concurrently, so there is an overlap between their methodological procedures. Nevertheless, each review has unique testing data and differing effect modifications from participants’ smoking history that necessitate conducting separate reviews for respiratory and cardiovascular health effects.

These two reviews will be living systematic reviews. Living systematic reviews are a recent innovation for conducting systematic reviews that incorporates evidence from studies as they are published [31, 32]. Living systematic reviews follow the established methods of conducting a systematic review, and in addition perform searches and publish updated reviews at pre-specified intervals. This enhancement overcomes the major issue of systematic reviews becoming outdated soon after publication [33].

Cochrane guidelines [32] specify three conditions to justify conducting a living systematic review: (1) a lack of high-quality studies or certainty about them, (2) the priority of the evidence for decision-making, and (3) emerging data that have a significant impact on the conclusions of prior reviews. Our review questions satisfy all the above conditions. First, there is a relatively limited number of human studies on the health effects of ENDS, contributing to the uncertainty on their clinical impact [27, 34, 35]. Second, decisions by clinicians and policy makers are often based on beliefs that are not supported by the available studies [36, see also 37 38]. Research evidence on the health risks of ENDS is a priority for decision-making because ENDS could represent an excellent tobacco harm reduction opportunity [34] if their long-term safety would be demonstrated. Finally, new evidence is being published that would be expected to impact existing knowledge. Indeed, the research output on ENDS has increased rapidly since 2018 [39, 40]. Our preliminary search of articles on ENDS in PubMed retrieved 1,332 articles published in 2019 and 1,565 articles in 2020 (search in Supplementary Materials). The living systematic review format is clearly in order.

The review team is composed of RO, the project leader, with extensive experience in conducting literature reviews and a substantial background in tobacco control, tobacco harm reduction, and ENDS. The research team is composed of four fellows. MQ, GRMLR, and DO each having literature review experience as first author of a narrative review, and RV who has substantial experience in literature reviews. MQ and GRMLR are clinicians, and DO has worked as a pharmacist. MQ has a background in tobacco control and tobacco harm reduction.

The following processes will be conducted to create the baseline reviews.

### Study Selection

Study designs selected for the reviews are randomized and non-randomized controlled trials and clinical trials, prospective and retrospective cohort studies, and case controlled studies. A grey literature search was conducted. Database searches were performed separately for cardiovascular and respiratory studies. Supplementary searches were conducted after the database search and after the full paper selection.

### Grey Literature Search

A search for papers on ENDS not published in peer reviewed journals was conducted on January 13, 2021 at the websites of 41 cardiovascular medical organizations and 53 respiratory medical organizations (see Supplementary Materials). No grey literature was located.

### Database search

Databases: Scopus, PUBMED and CENTRAL Cochrane Library

Keywords: ENDS keywords: “electronic cigarettes,” and “e-cigarettes”. Vapor or vapour were not used for keywords as these terms retrieve many chemical studies. Cardiovascular keywords: cardiovascular, heart, circulatory, arterial, and stroke. Respiratory keywords: lung, pulmonary, and respiratory.

Text fields searched were title and abstract in PUBMED; title, abstract and keyword in SCOPUS; and trials in Cochrane Library.

The search dates were 2010 through January 31, 2021. The start date of 2010 is the date of the publication of the first peer-reviewed research studies on ENDS.

The languages searched were English, French, Spanish, and studies in any language with an English abstract.

In compliance with PRISMA-P, an example of the search strategy is reported in Supplementary Materials.

The retrievals were entered into EndNote for bibliographic management. The paper retrievals were deduplicated. Cardiovascular retrievals were 374 and respiratory retrievals were 703.

The first exclusion of articles was performed on titles, and where a title was not sufficient for a determination, the abstract was reviewed. There were five categories of exclusion criteria. The first was article types: editorials, commentaries, letters, news articles, issue introductions, and reviews. The second was studies not peer reviewed including conference abstracts, unpublished clinical trials, and pre-print articles. The third was articles without an English abstract (except French or Spanish language). The fourth was studies exclusively on EVALI (e-cigarette or vaping product use associated lung injury). The fifth was on study design: in vitro, inhalation toxicology, biomarker, animal, cross-sectional studies, and studies conducted exclusively with youth.

The exclusion process was conducted independently by two reviewers and all discrepancies were resolved by discussion between the reviewers. One study in a non-included language was excluded. The count of studies after the title and abstract exclusion were 43 cardiovascular studies and 23 respiratory studies.

As a supplementary search, the reviews discussed in the introduction [15, 18-28] were searched independently by two reviewers for studies with the exclusion criteria above.

The second process of inclusion and exclusion was a full paper review. There were three inclusion criteria. One, studies were limited to the research designs of randomized controlled trials, non-randomized clinical studies, prospective or retrospective cohort studies, and case serries. Two, a study was required to have either a comparator of participants who smoked combustible tobacco (cigarettes) or a before and after testing of participants who had substituted ENDS for smoking. Third, the study had to report outcome data on cardiovascular or respiratory functioning or diseases. All three criteria had to met for a study to be included.

Review team members were trained and supervised on the inclusion criteria on 5 studies. The inclusion and exclusion of studies was conducted independently by two reviewers, and discrepancies were resolved by discussion. The reviewers achieved 95% agreement on study inclusion and exclusion. The project leader made the final disposition on studies that were questioned.

After this step the reference lists of all studies included were reviewed for additional studies and were citation chased in Google Scholar. A list of studies excluded during the full paper review is reported in the Supplemental Materials.

The search processes yielded 36 cardiovascular studies and 19 respiratory studies listed in Supplementary Materials. The list of the included studies for each review will be sent to two medical experts for examination to assure that no relevant studies have been missed. Any studies added will be reported in the review.

### Data extraction process

The data extraction process is being conducted with a data extraction form. The reviewers were trained with calibration exercises to ensure consistency. Data is being extracted independently by two reviewers, and then cross-checked by the reviewers for accuracy and completeness. Any discrepancies in data extraction will be rectified by discussion The data extraction items were drawn from inventories by JBI and the Cochrane Collaboration [41, 42]. The data extraction form is in Supplementary Materials. The categories include bibliographic details, population data, description of the intervention, respiratory or cardiovascular functioning or disease outcomes, data analysis, and study conclusions. If the published data is judged as insufficient or missing, the corresponding author will be sent an email with a request for additional details.

Two additional examinations of each study are being performed. The first is a check for internal discrepancies in the reporting of data [43] (form in Supplementary Materials). The second is a comparison of the study as it was conducted with its protocol or clinical trial registration where one is available (form in Supplementary Materials).

The completed data extraction forms will be submitted to the Systematic Review Data Repository at the Agency for Healthcare Research and Quality, an open access database.

Data from the studies will be reported in two ways. First, individual studies will be presented with a brief narrative description. Second, study tables will be constructed with items from the data extractions and the quality assessments (see following section).

### Quality Assessment and Risk of Bias

A quality assessment of each study is being conducted with the JBI quality assessment tool for its research design [45]. Reviewers were trained on the JBI quality assessment tools with an examination of its questions and a discussion of examples of quality issues. The quality assessment of the statistical analyses will be double checked by a reviewer (RO or RV) with training in biomedical statistics.

Bias is being assessed with observations of biases listed in the Oxford Centre for Evidence Based Medicine *Catalogue of Bias* [46] applicable to the study designs in the review. The review team prepared a set of prompting questions for each type of bias, and were briefed on common examples. The checklist of biases is in the Supplementary Materials.

The quality assessment and risk of bias observations are being made concurrently with the data extraction process. Two reviewers are independently performing the quality and bias assessments. Discrepancies will be resolved by discussion between the reviewers. If consensus is not reached, the final disposition will be made by the project leader. No studies will be excluded based on its quality assessment or risk of bias observations. Studies not conforming to JBI quality assessment items or with observations of biases will be reported in the study tables. These shortcomings will be specified in the data analysis and will be referenced in the discussion section of the reviews.

### Data analysis and synthesis

The synthesis will be a narrative synthesis. Due to the heterogeneity of the study populations and outcome measurements, we anticipate that a meta-analysis will be inappropriate. If sufficient studies are identified as comparable either during the baseline review or for updated reviews, we will develop an additional protocol for a meta-analysis and will add it to the review.

The narrative syntheses will have four components. The first will be findings organized by study design. The second will be a summary of descriptive statistics for participants and intervention characteristics. The third will group findings by the tests performed, physiological functions, and disease outcomes. The final synthesis will tally studies that have been assessed as demonstrating quality issues or biases.

Three sub-group analyses of testing and disease outcomes will be conducted for (1) concurrent users of cigarettes (dual users), (2) populations with prior respiratory/cardiovascular diseases, and (3) ENDS use of a duration of one year or longer.

Sensitivity analyses will be performed to explore the influence of risk of bias on the findings. One will be to rerun the third analysis by excluding all studies assessed at high risk of bias. A set of sensitivity analyses will be performed on groups of studies based on the following types of funding sources and author affiliations: ENDS/tobacco industry, pharmaceutical affiliations, and philanthropic or medical organization affiliations or funding. Publication bias will be assessed with a funnel plot for test results and disease outcomes.

The certainty evidence for each reviews’ findings will be evaluated with the Grading of Recommendations Assessment, Development and Evaluation (GRADE) [47, 48].

### Update plan

#### Updating the Study Search

A literature search will be conducted at three month intervals to retrieve newly published studies. Searches will be conducted with the same databases and keywords as the baseline review. Newly published papers will be checked for studies meeting the inclusion criteria. Searches will apply appropriate date limiters to include only those records added to the database subsequent to the last search, allowing for indexing lag time [31, 32]. Searches will be maintained in an EndNote library that will record all the search results over the lifetime of the review. Notes about each new search and the inclusion/exclusion decision for each study will be recorded. The newly included studies will be checked for references. The PRISMA diagram will be updated accordingly.

The first update search will be conducted on April 30 2021. Studies retrieved will be incorporated into the baseline review so that it will be up-to-date when published.

The updating search can result in three scenarios: 1) no new studies are identified (highly unlikely), 2) new studies are retrieved but the evidence has no impact on the review’s findings, or 3) new studies are found with evidence that significantly impacts the review’s findings. This third scenario will trigger an immediate updating of the review [31, 32]. In evaluating the impact of new evidence, we will consider whether it causes a change in the GRADE certainty rating or introduces previously unreported interventions, populations, serious adverse events, or other clinically meaningful findings [32, 49]. Any of these conditions will trigger conducting an updated review.

#### Updating the Review

Because a high volume of new studies is expected, we will update the review every six months if no studies prompt an earlier revision of the review. Narrative descriptions of the newly included studies and new study table listings will be produced merging them with the baseline review studies. All data analyses will be rerun with the procedures used for the baseline review. Conclusions and recommendations will be revised to reflect the addition of the new studies.

In addition, the search methods will be reviewed annually, or sooner if substantial changes occur that impact the search methodology, such as new search terms or sources. Possible changes to the frequency of the study searches will be considered. We will verify that the scope of the review (i.e. PICO components) is reflected in the inclusion of adequate thesaurus terms or changes to database search syntax. Other methodological aspects such as the use of technology enablers will also be reviewed annually.

#### Transition Out of Living Mode

The review will be transitioned out of living mode if the research question no longer meets all three criteria justifying the living approach. At that time we will examine article-level metrics, knowledge translation activities, and the output of new research studies. A practical factor that could result in the end of the living mode is reaching the end of our funding in September 2023. After this period, new funding will be sought to maintain the living mode. If funding is not acquired, we will ask if at least two members of the review team are available as volunteers for one year of updates followed with a final revision of the review. If both these strategies fail, the review will be transitioned into a traditional systematic review for final publication at the end of the project.

## Results

The goal of these reviews is to assemble all the available human studies on the respiratory and cardiovascular effects of ENDS for people who smoke. Furthermore, the reviews will assess the quality and potential biases of the studies to foreground the best available evidence. The reviews will identify those studies that demonstrate reporting bias so that the misrepresentation of ENDS health effects can be addressed in the contentious debate around tobacco harm reduction. The living systematic review methodology will keep the evidence current and complete as opposed to a static systematic review that quickly becomes out-of-date due to the rapid pace of publication of ENDS studies.

As of March 11, 2020 the literature search has been completed except for the review of the study selection by experts, with 36 studies included in the cardiovascular review and 19 studies in the respiratory review. Training on data extraction, quality assessment, and bias assessment processes has been completed. The data extraction process has just started, with 3 studies completed for the cardiovascular review and 1 study for the respiratory review. The target date for the completion of the reviews is July 2021.

## Discussion

Few protocols contain substantive planning for dissemination or knowledge translation activities beyond publication of the review in a peer-reviewed journal and conference presentations. Of course the reviews will be disseminated through these traditional avenues.

Pending acceptance, the projects’ protocol will be published on medRxiv and in the Journal of Medical Internet Research Protocols. The baseline and updated reviews will be published in a peer reviewed journal that agrees to work with the living format for updated editions of the review. The abstracts for the reviews will be translated into as many languages of possible. Announcements of these publications will be sent out on social media platforms (Twitter, Facebook, etc.).

In addition, we will write white papers to make the findings of the reviews accessible to clinicians and current or potential ENDS users, and policy briefs for policy makers. The reviews and white papers will be published for downloading on a dedicated website. The reviews, white papers, and policy briefs will be contributed as references to the relevant Wikipedia pages.

For clinicians, a white paper will spell out the treatment considerations of ENDS use drawn from both the cardiovascular and respiratory reviews. Because the large majority of physicians and healthcare providers hold erroneous beliefs about the health effects of nicotine itself [50-52] the white paper will include a section on nicotine. The white paper will be translated into as many languages as possible and sent to medical associations, distributed at conferences, and published on a website for downloading.

For current and potential ENDS users, a white paper with infographics will explain the health effects found in the reviews. The public also holds misperceptions about the health effects of nicotine [11, 53], so this white paper will have a section on nicotine. The white paper will be sent to the International Network of Nicotine Consumer Organizations, vapour product magazines, the Cochrane Consumer Network, Consumers United for Evidence-Based Healthcare, and patient advocacy organizations concerned with smoking-related diseases. We will explore producing short videos for YouTube and TED EX based on the white paper.

A policy brief on the potential public health effects of ENDS will be sent to national health departments and government legislators on health committees. The policy brief will be in one of their official languages and will be tailored to the country’s smoking prevalence and regulatory environment. We do not anticipate having the resources to write a policy brief for every country, so we will focus our initial efforts on countries that ban ENDS.

Our goal with these actions is to achieve a wide dissemination of the findings of the reviews to multiple audiences. Standard review publication practices would fail to reach the stakeholders who will benefit from having the evidence presented in a format that is accessible and readily understood.

The terrible toll of death and disease from cigarette smoking calls for every effort to stem the tobacco epidemic. The substitution of ENDS for cigarettes is one of the ways to potentially reduce the risks of smoking. Clinicians, and their patients who smoke need to understand the benefits of substituting ENDS for cigarettes. They also need to be aware of the risks because *research toward uncovering the risks of e-cigarette use is aligned with optimizing harm reduction* [54, p. 36]. Our living systematic reviews seek to highlight the best and most up-to-date evidence in this highly contentious and fast-moving field of research.

## Data Availability

Not applicable.

## Author Contributions

RO and RP conceptualized the reviews. RO conducted the database search and RV and DO conducted the grey literature search. RO, MQ, GRMLR, RV, and DO contributed to the manuscript. RO revised the manuscript. RP and GB reviewed an initial draft and the final manuscript for accuracy and completeness. All authors have read and approved the final manuscript.

## Author Declarations

RO is supported by a contract with ECLAT, Srl. ECLAT has received funding from the Foundation for a Smoke-Free World. She declares no conflicts of interest.

MQ has a grant from Foundation for a Smoke-Free World, outside the submitted work, for the project Pakistan Tobacco Economics Research and Dissemination. She declares no conflict of interest.

GRMLR, RV, DO, and GB declare no conflicts of interest.

RP has received has received lecture fees and research funding from Pfizer, GlaxoSmithKline, CV Therapeutics, NeuroSearch A/S, Sandoz, MSD, Boehringer Ingelheim, Novartis, Duska Therapeutics, and Forest Laboratories. He has served as a consultant for Pfizer, Global Health Alliance for treatment of tobacco dependence, CV Therapeutics, Boehringer Ingelheim, Novartis, Duska Therapeutics, ECITA (Electronic Cigarette Industry Trade Association, in the UK), Arbi Group Srl., and Health Diplomats. He has served on the Medical and Scientific Advisory Board (MSAB) of Cordex Pharma, Inc., CV Therapeutics, Duska Therapeutics Inc, Pfizer, and PharmaCielo. Lecture fees from a number of European EC industry and trade associations (including FIVAPE in France and FIESEL in Italy) were directly donated to vaper advocacy non-profit organizations. RP is the founder of the Center for Tobacco prevention and treatment (CPCT) at the University of Catania and the Center of Excellence for the acceleration of Harm Reduction (CoEHAR) at the same University, which has received support from Foundation for a Smoke Free World to conduct 8 independent investigator-initiated research projects on harm reduction. RP is currently involved in the following pro bono activities: scientific advisor for LIAF, Lega Italiana Anti Fumo (Italian acronym for Italian Anti-Smoking League), the Consumer Advocates for Smoke-free Alternatives (CASAA) and the International Network of Nicotine Consumers Organizations (INNCO); Chair of the European Technical Committee for standardization on “Requirements and test methods for emissions of electronic cigarettes” (CEN/TC 437; WG4).

