## Supplementary Materials for "Respiratory and Cardiovascular Health Effects of E-Cigarette Substitution: Protocol for Two Living Systematic Reviews"

A. PubMed bibliographic search for 2019 and 2020……………………………..2

B. Cardiovascular medical organizations grey literature search………………….3

C. Respiratory medical organization grey literature search…………….……….5

D. Sample search: PubMed respiratory…………………………………………...7

E. Full paper review exclusions – cardiovascular………………………..……8

F. Full paper review exclusions – respiratory……………………………………14

G. Cardiovascular studies included……………………………………..…….16

H. Respiratory studies included……………………………………..…………23

I. Data extraction form………………………………………...…………….27

J. Data discrepancies form……………………………………..……………30

K. Protocol discrepancies form……………………………………...…………31

L. Bias report…………………………………………………………..…………32

M. PRISMA-P 2015 Checklist……………………………………………………34

A. PubMed bibliographic search for 2019 and 2020

Field: title/abstract

Terms: e-cigarette OR “electronic cigarette” OR “electronic nicotine” OR vaping

Search conducted July 11, 2020

Retrivals

2019: 1284

2019 Epublished in 2018: 214

2019 Epublished in 2017: 6

Total published in 2019

1284 – (214 + 6) + 268 = 1,332

Search conducted January 19, 2021

2020: 1853

2020 Epublished in 2019: 268

2020 Epublished in 2018:20

Total published in 2020

1853 – (268 + 20) = 1565

#### B. Cardiovascular medical organizations grey literature search

| | |
|---|---|
| American Heart Organisation | https://www.heart.org/ |
| European Society of Cardiology | https://www.escardio.org/ |
| American College of Cardiology | https://www.acc.org/#sort=%40commonsortdate%20descending |
| Indian Association of Clinical Cardiologist | https://www.accindia.org/ |
| British Cardiovascular Society | https://www.britishcardiovascularsociety.org/ |
| British Junior Cardiologist Association | https://bjca.tv/ |
| Canadian Cardiovascular Society | https://ccs.ca/ |
| Cardiac Society of Australia & New Zealand | https://www.csanz.edu.au/ |
| Irish Cardiac Society | https://www.irishcardiacsociety.com/pages/default.asp |
| Emirates Cardiac Society | https://ecsociety.com/ |
| Caribbean Cardiac Society | https://cardiac.memberclicks.net/ |
| Scottish Cardiac Society | https://scottishcardiac.org/ |
| Cardiac Society of Nepal | http://www.csn.org.np/ |
| Pakistan Cardiac Society | http://www.pcs.org.pk/ |
| Adult Congenital Heart Association | https://www.achaheart.org/ |
| Gulf Heart Association | https://gulfheart.org/ |
| Philippines Heart Association | https://www.philheart.org/ |
| Italian Federation of Cardiologists | https://www.federcardio.it/en |
| Singapore Cardiac Society | https://www.singaporecardiac.org/ |
| Heart and Stroke Foundation of Canada | https://www.heartandstroke.ca/ |
| Heart Rhythm Society | https://www.hrsonline.org/ |
| Atrial Fibrillation Association | https://www.heartrhythmalliance.org/afa/uk/ |
| World Heart Federation | https://www.world-heart-federation.org/ |
| British Heart Foundation | https://www.bhf.org.uk/ |
| Royal College of Physicians | https://www.rcplondon.ac.uk/ |
| American College of Chest Physicians | https://www.chestnet.org/ |
| Cardiac Risk in the Young | https://www.c-r-y.org.uk/ |
| Thrombosis UK | https://thrombosisuk.org/ |
| Stroke Association | https://www.stroke.org.uk/ |
| American Stroke Association | https://www.stroke.org/ |
| World Stroke Organisation | https://www.world-stroke.org/ |
| European Stroke Organisation | https://eso-stroke.org/ |
| Singapore National Stroke Organisation | https://www.snsa.org.sg/ |
| Stroke Foundation | https://strokefoundation.org.au/ |
| Indian Stroke Association | http://www.stroke-india.org/ |
| Pacific Stroke Organisation | https://pacificstrokeassociation.org/ |
| Stroke Society of Australasia | https://www.strokesociety.com.au/ |
| Nordic Stroke Society | https://nordicstroke.org/ |

| | |
|---|---|
| Saudi Stroke Society | https://stroke.org.sa/ |
| Pakistan Stroke Society | http://pakstroke.com/ |
| Korean Stroke Society | https://www.stroke.or.kr:4454/eng/ |

C. Respiratory medical organizations grey literature search

1. American Association for Respiratory Care
   AARC.org
2. American College of Chest Physicians
   chestnet.org
3. American Lung Association
   https://www.lung.org/
4. American Respiratory Care Foundation
   ARCFoundation.org
5. American Thoracic Society
   thoracic.org
6. Argentine Association of Respiratory Medicine
   https://www.aamr.org.ar/
7. Asian Pacific Society of Respirology
   apsresp.org
8. Australian Lung Foundation
   lungfoundation.com.au
9. Australian & New Zealand Society of Respiratory Science
   anzsrs.org.au
10. Bangladesh Lung Foundation
    bdlungfoundation.com
11. Brazilian Society of Pulmonology and Physiology
    https://sbpt.org.br/portal/
12. BREATHE
    https://www.lung.ca/
13. British Thoracic Society
    brit-thoracic.org.uk
14. Chinese Association of Chest Physicians
    cacpchina.org/
15. Canadian Society of Respiratory Therapists
    https://www.csrt.com/
16. Canadian Thoracic Society
    lung.ca
17. Chinese Society of Respiratory Diseases
    csrd.org.cn
18. Chinese Thoracic Society
    ctschina.org/cn/
19. European Respiratory Society
    ersnet.org
20. European Society of Thoracic Surgeons
    https://www.ests.org/
21. Global Initiative for Asthma
    ginasthma.com
22. Global Initiative for Obstructive Lung Disease
    goldcopd.com

23. Ho Chi Minh City Respiratory Society
    hoihohaptphcm.org
24. Hong Kong Lung Foundation
    hklf.org
25. Hong Kong & Macau Chapter of the American College of Chest Physicians
    fmshk.org
26. Hong Kong Tuberculosis, Chest & Heart Disease Association
    antitb.org.hk
27. Hong Kong Thoracic Society
    hkts.hk
28. Indian Chest Society
    indianchestsociety.com/
29. International Primary Care Respiratory Group
    theipcrg.org
30. Indonesian Society of Respirology
    klikpdpi.com
31. International Society for Respiratory Diseases
    isrd.org
32. International Union Against Tuberculosis & Lung Disease
    theunion.org
33. Japanese Respiratory Society
    jrs.or.jp
34. Korean Academy of Tuberculosis and Respiratory Disease
    lungkorea.org/
35. Latin American Thoracic Association
    alatorax.org
36. Lung Foundation Australia
    lungfoundation.com.au
37. Malaysian Thoracic Society
    mts.org.my
38. Nigerian Thoracic Society
    http://nigerianthoracicsociety.org/
39. Pan African Thoracic Society
    https://panafricanthoracic.org/
40. Philippine College of Chest Physicians
    philchest.org
41. Respiratory Care Indonesia
    respina.org
42. Sri Lanka College of Pulmonologists
    copsl.lk
43. Saudi Thoracic Society
    saudithoracic.com
44. Singapore Thoracic Society
    thoracic.sg

D. Sample search: PubMed respiratory

Search: **(((respiratory[Title/Abstract]) OR (lung[Title/Abstract])) OR (pulmonary[Title/Abstract])) AND ("e-cigarette"[Title/Abstract])** Filters: **from 2010 - 2021** Sort by: **Most Recent**

Search: **(((respiratory[Title/Abstract]) OR (lung[Title/Abstract])) OR (pulmonary[Title/Abstract])) AND ("electronic nicotine"[Title/Abstract])** Sort by: **Most Recent (first study 2013)**

E. Full paper review exclusions – cardiovascular

TOTAL: 38

Not an included study design - 5

Alzahrani, T., I. Pena, N. Temesgen and S. A. Glantz (2018). "Association Between Electronic Cigarette Use and Myocardial Infarction." American Journal of Preventive Medicine **55**(4): 455-461.

Boas, Z., P. Gupta, R. S. Moheimani, M. Bhetraratana, F. Yin, K. M. Peters, J. Gornbein, J. A. Araujo, J. Czernin and H. R. Middlekauff (2017). "Activation of the “Splenocardiac Axis” by electronic and tobacco cigarettes in otherwise healthy young adults." Physiological Reports **5**(17).

Ikonomidis, I., D. Vlastos, K. Kourea, G. Kostelli, M. Varoudi, G. Pavlidis, P. Efentakis, H. Triantafyllidi, J. Parissis, I. Andreadou, E. Iliodromitis and J. Lekakis (2018). "Electronic Cigarette Smoking Increases Arterial Stiffness and Oxidative Stress to a Lesser Extent Than a Single Conventional Cigarette: An Acute and Chronic Study." Circulation **137**(3): 303-306.

Kim, C. Y., Y. J. Paek, H. G. Seo, Y. S. Cheong, C. M. Lee, S. M. Park, D. W. Park and K. Lee (2020). "Dual use of electronic and conventional cigarettes is associated with higher cardiovascular risk factors in Korean men." Scientific Reports **10**(1).

Vlachopoulos, C., N. Ioakeimidis, M. Abdelrasoul, D. Terentes-Printzios, C. Georgakopoulos, P. Pietri, C. Stefanadis and D. Tousoulis (2016). "Electronic Cigarette Smoking Increases Aortic Stiffness and Blood Pressure in Young Smokers." J Am Coll Cardiol **67**(23): 2802-2803.

No comparator to smoking – 25

Antoniewicz, L., J. A. Bosson, J. Kuhl, S. M. Abdel-Halim, A. Kiessling, F. Mobarrez and M. Lundbäck (2016). "Electronic cigarettes increase endothelial progenitor cells in the blood of healthy volunteers." Atherosclerosis **255**: 179-185.

Antoniewicz, L., A. Brynedal, L. Hedman, M. Lundbäck and J. A. Bosson (2019). "Acute

Effects of Electronic Cigarette Inhalation on the Vasculature and the Conducting Airways." Cardiovascular Toxicology **19**(5): 441-450.

Arefalk, G., K. Hambraeus, L. Lind, K. Michaelsson, B. Lindahl and J. Sundstrom (2014). "Discontinuation of smokeless tobacco and mortality risk after myocardial infarction." Circulation **130**(4): 325-332.

Caporale, A., M. C. Langham, W. Guo, A. Johncola, S. Chatterjee and F. W. Wehrli (2019). "Acute Effects of Electronic Cigarette Aerosol Inhalation on Vascular Function Detected at Quantitative MRI." Radiology **293**(1): 97-106.

Chaumont, M., B. De Becker, W. Zaher, A. Culié, G. Deprez, C. Mélot, F. Reyé, P. Van Antwerpen, C. Delporte, N. Debbas, K. Z. Boudjeltia and P. Van De Borne (2018). "Differential Effects of E-Cigarette on Microvascular Endothelial Function, Arterial Stiffness and Oxidative Stress: A Randomized Crossover Trial." Scientific Reports **8**(1).

Chaumont, M., V. Tagliatti, E. M. Channan, J. M. Colet, A. Bernard, S. Morra, G. Deprez, A. van Muylem, N. Debbas, T. Schaefer, V. Faoro and P. van de Borne (2020). "Short halt in vaping modifies cardiorespiratory parameters and urine metabolome: A randomized trial." American Journal of Physiology - Lung Cellular and Molecular Physiology **318**(2): L331-L344.

Cossio, R., Z. A. Cerra and H. Tanaka (2020). "Vascular effects of a single bout of electronic cigarette use." Clinical and Experimental Pharmacology and Physiology **47**(1): 3-6.

Goniewicz, M. L., M. Gawron, D. M. Smith, M. Peng, P. Jacob, 3rd and N. L. Benowitz (2017). "Exposure to Nicotine and Selected Toxicants in Cigarette Smokers Who Switched to Electronic Cigarettes: A Longitudinal Within-Subjects Observational Study." Nicotine Tob Res **19**(2): 160-167.

Gonzalez, J. E. and W. H. Cooke (2021). "Acute effects of electronic cigarettes on arterial pressure and peripheral sympathetic activity in young nonsmokers." Am J Physiol Heart Circ Physiol **320**(1): H248-h255.

Hajek, P., A. Phillips-Waller, D. Przulj, F. Pesola, K. Myers Smith, N. Bisal, J. Li, S. Parrott, P. Sasieni, L. Dawkins, L. Ross, M. Goniewicz, Q. Wu and H. J. McRobbie (2019). "A

Randomized Trial of E-Cigarettes versus Nicotine-Replacement Therapy." N Engl J Med **380**(7): 629-637.

Hiler, M., N. Karaoghlanian, S. Talih, S. Maloney, A. Breland, A. Shihadeh and T. Eissenberg (2020). "Effects of electronic cigarette heating coil resistance and liquid nicotine concentration on user nicotine delivery, heart rate, subjective effects, puff topography, and liquid consumption." Experimental and Clinical Psychopharmacology **28**(5): 527-539.

Hughes, J. R., E. N. Peters, P. W. Callas, C. Peasley-Miklus, E. Oga, J. F. Etter and N. Morley (2020). "Withdrawal Symptoms from E-Cigarette Abstinence among Adult Never-Smokers: A Pilot Experimental Study." Nicotine and Tobacco Research **22**(5): 740-746.

Hughes, J. R., E. N. Peters, P. W. Callas, C. Peasley-Miklus, E. Oga, J. F. Etter and N. Morley (2020). "Withdrawal Symptoms from E-Cigarette Abstinence among Former Smokers: A Pre-Post Clinical Trial." Nicotine and Tobacco Research **22**(5): 734-739.

Kinoshita, M., R. M. Herges, D. O. Hodge, L. Friedman, N. M. Ammash, C. J. Bruce, V. Somers, J. F. Malouf, J. Askelin, J. A. Gilles, B. J. Gersh and P. A. Friedman (2009). "Role of smoking in the recurrence of atrial arrhythmias after cardioversion." Am J Cardiol **104**(5): 678-682.

Lee, M. S., V. W. Rees, P. Koutrakis, J. M. Wolfson, Y. S. Son, J. Lawrence and D. C. Christiani (2019). "Cardiac Autonomic Effects of Secondhand Exposure to Nicotine from Electronic Cigarettes: An Exploratory Study." Environ Epidemiol **3**(1).

McClelland, M. L., C. S. Sesoko, D. A. MacDonald and L. M. Davis (2020). "Effects on vital signs after twenty minutes of vaping compared to people exposed to second-hand vapor." Advances in respiratory medicine **88**(6): 504-514.

Moheimani, R. S., M. Bhetraratana, K. M. Peters, B. K. Yang, F. Yin, J. Gornbein, J. A. Araujo and H. R. Middlekauff (2017). "Sympathomimetic effects of acute e-cigarette use: Role of nicotine and non-nicotine constituents." Journal of the American Heart Association **6**(9).

Nadruz, W., Jr., B. Claggett, A. Goncalves, G. Querejeta-Roca, M. M. Fernandes-Silva, A. M. Shah, S. Cheng, H. Tanaka, G. Heiss, D. W. Kitzman and S. D. Solomon (2016). "Smoking and

Cardiac Structure and Function in the Elderly: The ARIC Study (Atherosclerosis Risk in Communities)." Circ Cardiovasc Imaging **9**(9): e004950.

Polosa, R., F. Cibella, P. Caponnetto, M. Maglia, U. Prosperini, C. Russo and D. Tashkin (2017). "Health impact of E-cigarettes: A prospective 3.5-year study of regular daily users who have never smoked." Scientific Reports **7**(1).

Pywell, M. J., M. Wordsworth, R. M. Kwasnicki, P. Chadha, S. Hettiaratchy and T. Halsey (2018). "The Effect of Electronic Cigarettes on Hand Microcirculation." J Hand Surg Am **43**(5): 432-438.

Rüther, T., D. Hagedorn, K. Schiela, T. Schettgen, H. Osiander-Fuchs and W. Schober (2018). "Nicotine delivery efficiency of first- and second-generation e-cigarettes and its impact on relief of craving during the acute phase of use." International Journal of Hygiene and Environmental Health **221**(2): 191-198.

Spindle, T. R., M. M. Hiler, A. B. Breland, N. V. Karaoghlanian, A. L. Shihadeh and T. Eissenberg (2017). "The Influence of a Mouthpiece-Based Topography Measurement Device on Electronic Cigarette User's Plasma Nicotine Concentration, Heart Rate, and Subjective Effects Under Directed and Ad Libitum Use Conditions." Nicotine & tobacco research : official journal of the Society for Research on Nicotine and Tobacco **19**(4): 469-476.

St.Helen, G., C. Havel, D. A. Dempsey, P. Jacob, III and N. L. Benowitz (2016). "Nicotine delivery, retention and pharmacokinetics from various electronic cigarettes." Addiction **111**(3): 535-544.

Wallenfeldt, K., J. Hulthe, L. Bokemark, J. Wikstrand and B. Fagerberg (2001). "Carotid and femoral atherosclerosis, cardiovascular risk factors and C-reactive protein in relation to smokeless tobacco use or smoking in 58-year-old men." J Intern Med **250**(6): 492-501.

Yatsuya, H., A. R. Folsom and A. Investigators (2010). "Risk of incident cardiovascular disease among users of smokeless tobacco in the Atherosclerosis Risk in Communities (ARIC) study." Am J Epidemiol **172**(5): 600-605.

No data on cardiovascular functions or diseases – 7

Bullen, C., C. Howe, M. Laugesen, H. McRobbie, V. Parag, J. Williman and N. Walker (2013). "Electronic cigarettes for smoking cessation: a randomised controlled trial." Lancet **382**(9905): 1629-1637.

Czogała, J., M. Cholewiński, A. Kutek and W. Zielińska-Danch (2012). "[Evaluation of changes in hemodynamic parameters after the use of electronic nicotine delivery systems among regular cigarette smokers]." Przegląd lekarski **69**(10): 841-845.

Flouris, A. D., K. P. Poulianiti, M. S. Chorti, A. Z. Jamurtas, D. Kouretas, E. O. Owolabi, M. N. Tzatzarakis, A. M. Tsatsakis and Y. Koutedakis (2012). "Acute effects of electronic and tobacco cigarette smoking on complete blood count." Food Chem Toxicol **50**(10): 3600-3603.

Hébert-Losier, A., K. B. Filion, S. B. Windle and M. J. Eisenberg (2020). "A Randomized Controlled Trial Evaluating the Efficacy of E-Cigarette Use for Smoking Cessation in the General Population: E3 Trial Design." CJC Open **2**(3): 168-175.

Hecht, S. S., S. G. Carmella, D. Kotandeniya, M. E. Pillsbury, M. Chen, B. W. Ransom, R. I. Vogel, E. Thompson, S. E. Murphy and D. K. Hatsukami (2015). "Evaluation of toxicant and carcinogen metabolites in the urine of e-cigarette users versus cigarette smokers." Nicotine Tob Res **17**(6): 704-709.

Manzoli, L., C. La Vecchia, M. E. Flacco, L. Capasso, V. Simonetti, S. Boccia, A. Di Baldassarre, P. Villari, A. Mezzetti and G. Cicolini (2013). "Multicentric cohort study on the long-term efficacy and safety of electronic cigarettes: Study design and methodology." BMC Public Health **13**(1).

Wagener, T. L., E. L. Floyd, I. Stepanov, L. M. Driskill, S. G. Frank, E. Meier, E. L. Leavens, A. P. Tackett, N. Molina and L. Queimado (2017). "Have combustible cigarettes met their match? The nicotine delivery profiles and harmful constituent exposures of second-generation and third-generation electronic cigarette users." Tobacco Control **26**(e1): e23-e28.

Other – not vaping

MacLean, R. R., R. Gueorguieva, E. E. DeVito, M. R. Peltier, S. Parida and M. Sofuoglu (2020). "The Effects of Inhaled Flavors on Intravenous Nicotine." Experimental and Clinical Psychopharmacology.

F. Full paper review exclusions - respiratory

Aherrera, A., A. Aravindakshan, S. Jarmul, P. Olmedo, R. Chen, J. E. Cohen, A. Navas-Acien and A. M. Rule (2020). "E-cigarette use behaviors and device characteristics of daily exclusive e-cigarette users in Maryland: Implications for product toxicity." Tob Induc Dis **18**: 93. No comparator.

Antoniewicz, L., A. Brynedal, L. Hedman, M. Lundbäck and J. A. Bosson (2019). "Acute Effects of Electronic Cigarette Inhalation on the Vasculature and the Conducting Airways." Cardiovascular Toxicology **19**(5): 441-450. No comparator.

Caponnetto, P., D. Campagna, F. Cibella, J. B. Morjaria, M. Caruso, C. Russo and R. Polosa (2013). "EffiCiency and Safety of an eLectronic cigAreTte (ECLAT) as tobacco cigarettes substitute: a prospective 12-month randomized control design study." PLoS One **8**(6): e66317. No respiratory outcomes.

Ferrari, M., A. Zanasi, E. Nardi, A. M. Morselli Labate, P. Ceriana, A. Balestrino, L. Pisani, N. Corcione and S. Nava (2015). "Short-term effects of a nicotine-free e-cigarette compared to a traditional cigarette in smokers and non-smokers." BMC Pulmonary Medicine **15**(1). No comparator.

Kizhakke Puliyakote, A. S., A. R. Elliott, R. C. Sá, K. M. Anderson, L. E. Crotty Alexander and S. R. Hopkins (2020). "Vaping Disrupts Ventilation-Perfusion Matching in Asymptomatic Users." J Appl Physiol. No comparator.

Lappas, A. S., A. S. Tzortzi, E. M. Konstantinidi, S. I. Teloniatis, C. K. Tzavara, S. A. Gennimata, N. G. Koulouris and P. K. Behrakis (2018). "Short-term respiratory effects of e-cigarettes in healthy individuals and smokers with asthma." Respirology **23**(3): 291-297. No comparator.

Lee, S. M., R. Tenney, A. W. Wallace and M. Arjomandi (2018). "E-cigarettes versus nicotine patches for perioperative smoking cessation: a pilot randomized trial." PeerJ **6**: e5609 No comparator.

Lucchiari, C., M. Masiero, K. Mazzocco, G. Veronesi, P. Maisonneuve, C. Jemos, E. Omodeo Salè, S. Spina, R. Bertolotti and G. Pravettoni (2020). "Benefits of e-cigarettes in smoking reduction and in pulmonary health among chronic smokers undergoing a lung cancer screening program at 6 months." Addictive Behaviors **103**.

Meo, S. A., M. A. Ansary, F. R. Barayan, A. S. Almusallam, A. M. Almehaid, N. S. Alarifi, T. A. Alsohaibani and I. Zia (2019). "Electronic Cigarettes: Impact on Lung Function and Fractional Exhaled Nitric Oxide Among Healthy Adults." American Journal of Men's Health **13**(1). No comparator.

Polosa, R., F. Cibella, P. Caponnetto, M. Maglia, U. Prosperini, C. Russo and D. Tashkin (2017). "Health impact of E-cigarettes: A prospective 3.5-year study of regular daily users who have never smoked." Scientific Reports **7**(1). No comparator.

Xie, W., H. Kathuria, P. Galiatsatos, M. J. Blaha, N. M. Hamburg, R. M. Robertson, A. Bhatnagar, E. J. Benjamin and A. C. Stokes (2020). "Association of Electronic Cigarette Use With Incident Respiratory Conditions Among US Adults From 2013 to 2018." JAMA Netw Open **3**(11): e2020816. Excluded study design.

Xie, Z., D. J. Ossip, I. Rahman and D. Li (2020). "Use of Electronic Cigarettes and Self-Reported Chronic Obstructive Pulmonary Disease Diagnosis in Adults." Nicotine and Tobacco Research **22**(7): 1155-1161. Excluded study design.

G. Cardiovascular studies included

Arastoo, S., Haptonstall, K. P., Choroomi, Y., Moheimani, R., Nguyen, K., Tran, E., Gornbein, J., & Middlekauff, H. R. (2020). Acute and chronic sympathomimetic effects of e-cigarette and tobacco cigarette smoking: Role of nicotine and non-nicotine constituents. *American Journal of Physiology - Heart and Circulatory Physiology, 319*(2), H262-H270. https://doi.org/10.1152/ajpheart.00192.2020

Benowitz, N. L., St.Helen, G., Nardone, N., Addo, N., Zhang, J., Harvanko, A. M., Calfee, C. S., & Jacob, P. (2020). Twenty-Four-Hour Cardiovascular Effects of Electronic Cigarettes Compared With Cigarette Smoking in Dual Users. *Journal of the American Heart Association, 9*(23). https://doi.org/10.1161/jaha.120.017317

Biondi-Zoccai, G., Sciarretta, S., Bullen, C., Nocella, C., Violi, F., Loffredo, L., Pignatelli, P., Perri, L., Peruzzi, M., Marullo, A. G. M., De Falco, E., Chimenti, I., Cammisotto, V., Valenti, V., Coluzzi, F., Cavarretta, E., Carrizzo, A., Prati, F., Carnevale, R., & Frati, G. (2019, Mar 19). Acute Effects of Heat-Not-Burn, Electronic Vaping, and Traditional Tobacco Combustion Cigarettes: The Sapienza University of Rome-Vascular Assessment of Proatherosclerotic Effects of Smoking ( SUR - VAPES ) 2 Randomized Trial. *J Am Heart Assoc, 8*(6), e010455. https://doi.org/10.1161/JAHA.118.010455

Caponnetto, P., Campagna, D., Cibella, F., Morjaria, J. B., Caruso, M., Russo, C., & Polosa, R. (2013). EffiCiency and Safety of an eLectronic cigAreTte (ECLAT) as tobacco cigarettes substitute: a prospective 12-month randomized control design study. *PLoS ONE, 8*(6), e66317. https://doi.org/10.1371/journal.pone.0066317

Carnevale, R., Sciarretta, S., Violi, F., Nocella, C., Loffredo, L., Perri, L., Peruzzi, M., Marullo, A. G., De Falco, E., Chimenti, I., Valenti, V., Biondi-Zoccai, G., & Frati, G. (2016, Sep). Acute Impact of Tobacco vs Electronic Cigarette Smoking on Oxidative Stress and Vascular Function. *Chest, 150*(3), 606-612. https://doi.org/10.1016/j.chest.2016.04.012

Chaumont, M., van de Borne, P., Bernard, A., Van Muylem, A., Deprez, G., Ullmo, J., Starczewska, E., Briki, R., de Hemptinne, Q., Zaher, W., & Debbas, N. (2019). Fourth generation e-cigarette vaping induces transient lung inflammation and gas exchange disturbances: Results from two randomized clinical trials. *American Journal of Physiology - Lung Cellular and Molecular Physiology, 316*(5), L705-L719. https://doi.org/10.1152/ajplung.00492.2018

Cioe, P. A., Mercurio, A. N., Lechner, W., Costantino, C. C., Tidey, J. W., Eissenberg, T., & Kahler, C. W. (2020). A pilot study to examine the acceptability and health effects of electronic cigarettes in HIV-positive smokers. *Drug and Alcohol Dependence, 206*. https://doi.org/10.1016/j.drugalcdep.2019.107678

Cravo, A. S., Bush, J., Sharma, G., Savioz, R., Martin, C., Craige, S., & Walele, T. (2016, Nov 15). A randomised, parallel group study to evaluate the safety profile of an electronic vapour product over 12 weeks. *Regul Toxicol Pharmacol, 81 Suppl 1*, S1-S14. https://doi.org/10.1016/j.yrtph.2016.10.003

D'Ruiz, C. D., O'Connell, G., Graff, D. W., & Yan, X. S. (2017). Measurement of cardiovascular and pulmonary function endpoints and other physiological effects following partial or complete substitution of cigarettes with electronic cigarettes in adult smokers. *Regulatory Toxicology and Pharmacology, 87*, 36-53. https://doi.org/10.1016/j.yrtph.2017.05.002

Eissenberg, T. (2010, Feb). Electronic nicotine delivery devices: ineffective nicotine delivery and craving suppression after acute administration. *Tob Control, 19*(1), 87-88. https://doi.org/10.1136/tc.2009.033498

Farsalinos, K., Cibella, F., Caponnetto, P., Campagna, D., Morjaria, J. B., Battaglia, E., Caruso, M., Russo, C., & Polosa, R. (2016). Effect of continuous smoking reduction and abstinence on blood pressure and heart rate in smokers switching to electronic cigarettes. *Internal and Emergency Medicine, 11*(1), 85-94. https://doi.org/10.1007/s11739-015-1361-y

Farsalinos, K. E., Tsiapras, D., Kyrzopoulos, S., Savvopoulou, M., & Voudris, V. (2014, Jun 23). Acute effects of using an electronic nicotine-delivery device (electronic cigarette) on myocardial function: comparison with the effects of regular cigarettes. *BMC Cardiovasc Disord*, *14*, 78. https://doi.org/10.1186/1471-2261-14-78

Fetterman, J. L., Keith, R. J., Palmisano, J. N., McGlasson, K. L., Weisbrod, R. M., Majid, S., Bastin, R., Stathos, M. M., Stokes, A. C., Robertson, R. M., Bhatnagar, A., & Hamburg, N. M. (2020). Alterations in Vascular Function Associated With the Use of Combustible and Electronic Cigarettes. *Journal of the American Heart Association, 9*(9), e014570. https://doi.org/10.1161/JAHA.119.014570

Franzen, K. F., Willig, J., Cayo Talavera, S., Meusel, M., Sayk, F., Reppel, M., Dalhoff, K., Mortensen, K., & Droemann, D. (2018). E-cigarettes and cigarettes worsen peripheral and central hemodynamics as well as arterial stiffness: A randomized, double-blinded pilot study. *Vascular Medicine, 23*(5), 419-425. https://doi.org/10.1177/1358863X18779694

George, J., Hussain, M., Vadiveloo, T., Ireland, S., Hopkinson, P., Struthers, A. D., Donnan, P. T., Khan, F., & Lang, C. C. (2019). Cardiovascular Effects of Switching From Tobacco Cigarettes to Electronic Cigarettes. *Journal of the American College of Cardiology, 74*(25), 3112-3120. https://doi.org/10.1016/j.jacc.2019.09.067

Haptonstall, K. P., Choroomi, Y., Moheimani, R., Nguyen, K., Tran, E., Lakhani, K., Ruedisueli, I., Gornbein, J., & Middlekauff, H. R. (2020). Differential effects of tobacco cigarettes and electronic cigarettes on endothelial function in healthy young people. *American Journal of Physiology - Heart and Circulatory Physiology, 319*(3), H547-H556. https://doi.org/10.1152/ajpheart.00307.2020

Hiler, M., Breland, A., Spindle, T., Maloney, S., Lipato, T., Karaoghlanian, N., Shihadeh, A., Lopez, A., Ramôa, C., & Eissenberg, T. (2017). Electronic cigarette user plasma nicotine concentration, puff topography, heart rate, and subjective effects: Influence of liquid nicotine concentration and user experience. *Experimental and Clinical Psychopharmacology, 25*(5), 380-392. https://doi.org/10.1037/pha0000140

Ikonomidis, I., Katogiannis, K., Kostelli, G., Kourea, K., Kyriakou, E., Kypraiou, A., Tsoumani, M., Andreadou, I., Lambadiari, V., Plotas, P., Thymis, I., & Tsantes, A. E. (2020). Effects of electronic cigarette on platelet and vascular function after four months of use. *Food and Chemical Toxicology, 141*. https://doi.org/10.1016/j.fct.2020.111389

Ip, M., Diamantakos, E., Haptonstall, K., Choroomi, Y., Moheimani, R. S., Nguyen, K. H., Tran, E., Gornbein, J., & Middlekauff, H. R. (2020). Tobacco and electronic cigarettes adversely impact ECG indexes of ventricular repolarization: Implication for sudden death risk. *American Journal of Physiology - Heart and Circulatory Physiology, 318*(5), H1176-H1184. https://doi.org/10.1152/AJPHEART.00738.2019

Kerr, D. M. I., Brooksbank, K. J. M., Taylor, R. G., Pinel, K., Rios, F. J., Touyz, R. M., & Delles, C. (2019, Jan). Acute effects of electronic and tobacco cigarettes on vascular and respiratory function in healthy volunteers: a cross-over study. *J Hypertens, 37*(1), 154-166. https://doi.org/10.1097/HJH.0000000000001890

Kuntic, M., Oelze, M., Steven, S., Kröller-Schön, S., Stamm, P., Kalinovic, S., Frenis, K., Vujacic-Mirski, K., Jimenez, M. T. B., Kvandova, M., Filippou, K., Al Zuabi, A., Brückl, V., Hahad, O., Daub, S., Varveri, F., Gori, T., Huesmann, R., Hoffmann, T., Schmidt, F. P., Keaney, J. F., Jr., Daiber, A., & Münzel, T. (2020). Short-term e-cigarette vapour exposure causes vascular oxidative stress and dysfunction: Evidence for a close connection to brain damage and a key role of the phagocytic NADPH oxidase (NOX-2). *European Heart Journal, 41*(26), 2472-2483A. https://doi.org/10.1093/eurheartj/ehz772

Manzoli, L., Flacco, M. E., Fiore, M., La Vecchia, C., Marzuillo, C., Gualano, M. R., Liguori, G., Cicolini, G., Capasso, L., D'Amario, C., Boccia, S., Siliquini, R., Ricciardi, W., & Villari, P. (2015). Electronic cigarettes efficacy and safety at 12 months: Cohort study.

*PLoS ONE, 10*(6), Article e0129443. https://doi.org/10.1371/journal.pone.0129443

Mastrangeli, S., Carnevale, R., Cavarretta, E., Sciarretta, S., Peruzzi, M., Marullo, A. G. M., De Falco, E., Chimenti, I., Valenti, V., Bullen, C., Roever, L., Frati, G., & Biondi-Zoccai, G. (2018). Predictors of oxidative stress and vascular function in an experimental study of tobacco versus electronic cigarettes: A post hoc analysis of the SUR-VAPES 1 Study. *Tob Induc Dis, 16*, 18. https://doi.org/10.18332/tid/89975

Nides, M. A., Leischow, S. J., Bhatter, M., & Simmons, M. (2014, Mar). Nicotine blood levels and short-term smoking reduction with an electronic nicotine delivery system. *Am J Health Behav, 38*(2), 265-274. https://doi.org/10.5993/AJHB.38.2.12

Nocella, C., Biondi-Zoccai, G., Sciarretta, S., Peruzzi, M., Pagano, F., Loffredo, L., Pignatelli, P., Bullen, C., Frati, G., & Carnevale, R. (2018). Impact of Tobacco Versus Electronic Cigarette Smoking on Platelet Function. *American Journal of Cardiology, 122*(9), 1477-1481. https://doi.org/10.1016/j.amjcard.2018.07.029

Osibogun, O., Bursac, Z., McKee, M., Li, T., & Maziak, W. (2020). Cessation outcomes in adult dual users of e-cigarettes and cigarettes: the Population Assessment of Tobacco and Health cohort study, USA, 2013–2016. *International Journal of Public Health, 65*(6), 923-936. https://doi.org/10.1007/s00038-020-01436-w

Polosa, R., Morjaria, J. B., Caponnetto, P., Battaglia, E., Russo, C., Ciampi, C., Adams, G., & Bruno, C. M. (2016). Blood pressure control in smokers with arterial hypertension who switched to electronic cigarettes. *International Journal of Environmental Research and Public Health, 13*(11), 1123.

Sumartiningsih, S., Lin, H. F., & Lin, J. C. (2019). Cigarette smoking blunts exercise-induced heart rate response among young adult male smokers. *International Journal of Environmental Research and Public Health, 16*(6), Article 1032. https://doi.org/10.3390/ijerph16061032

Szołtysek-Bołdys, I., Sobczak, A., Zielińska-Danch, W., Bartoń, A., Koszowski, B., & Kośmider, L. (2014). Influence of inhaled nicotine source on arterial stiffness. *Przegląd lekarski, 71*(11), 572-575. https://www.scopus.com/inward/record.uri?eid=2-s2.0-84927170342&partnerID=40&md5=9182a1ae1acf30a07dfd6aa30a2ee1f1

van Staden, S. R., Groenewald, M., Engelbrecht, R., Becker, P. J., & Hazelhurst, L. T. (2013, Sep 30). Carboxyhaemoglobin levels, health and lifestyle perceptions in smokers converting from tobacco cigarettes to electronic cigarettes. *S Afr Med J, 103*(11), 865-868. https://doi.org/10.7196/samj.6887

Vansickel, A. R., Cobb, C. O., Weaver, M. F., & Eissenberg, T. E. (2010, Aug). A clinical laboratory model for evaluating the acute effects of electronic "cigarettes": nicotine delivery profile and cardiovascular and subjective effects. *Cancer Epidemiol Biomarkers Prev, 19*(8), 1945-1953. https://doi.org/10.1158/1055-9965.EPI-10-0288

Veldheer, S., Yingst, J., Midya, V., Hummer, B., Lester, C., Krebs, N., Hrabovsky, S., Wilhelm, A., Liao, J., Yen, M. S., Cobb, C., Eissenberg, T., & Foulds, J. (2019, Apr). Pulmonary and other health effects of electronic cigarette use among adult smokers participating in a randomized controlled smoking reduction trial. *Addict Behav, 91*, 95-101. https://doi.org/10.1016/j.addbeh.2018.10.041

Walele, T., Bush, J., Koch, A., Savioz, R., Martin, C., & O'Connell, G. (2018). Evaluation of the safety profile of an electronic vapour product used for two years by smokers in a real-life setting. *Regulatory Toxicology and Pharmacology, 92*, 226-238. https://doi.org/10.1016/j.yrtph.2017.12.010

Walele, T., Sharma, G., Savioz, R., Martin, C., & Williams, J. (2016, Feb). A randomised, crossover study on an electronic vapour product, a nicotine inhalator and a conventional cigarette. Part B: Safety and subjective effects. *Regul Toxicol Pharmacol, 74*, 193-199. https://doi.org/10.1016/j.yrtph.2015.12.004

Walker, N., Parag, V., Verbiest, M., Laking, G., Laugesen, M., & Bullen, C. (2020). Nicotine patches used in combination with e-cigarettes (with and without nicotine) for smoking cessation: a pragmatic, randomised trial. *The Lancet Respiratory Medicine*, *8*(1), 54-64. https://doi.org/10.1016/S2213-2600(19)30269-3

Yan, X. S., & D'Ruiz, C. (2015, Feb). Effects of using electronic cigarettes on nicotine delivery and cardiovascular function in comparison with regular cigarettes. *Regul Toxicol Pharmacol*, *71*(1), 24-34. https://doi.org/10.1016/j.yrtph.2014.11.004

H. Respiratory studies included

Barna, S., Rózsa, D., Varga, J., Fodor, A., Szilasi, M., Galuska, L., & Garai, I. (2019). First comparative results about the direct effect of traditional cigarette and e-cigarette smoking on lung alveolocapillary membrane using dynamic ventilation scintigraphy. *Nuclear Medicine Communications, 40*(2), 153-158. https://doi.org/10.1097/MNM.0000000000000957

Bowler, R. P., Hansel, N. N., Jacobson, S., Graham Barr, R., Make, B. J., Han, M. L. K., O'Neal, W. K., Oelsner, E. C., Casaburi, R., Barjaktarevic, I., Cooper, C., Foreman, M., Wise, R. A., DeMeo, D. L., Silverman, E. K., Bailey, W., Harrington, K. F., Woodruff, P. G., Drummond, M. B., for, C., & Investigators, S. (2017). Electronic Cigarette Use in US Adults at Risk for or with COPD: Analysis from Two Observational Cohorts. *Journal of General Internal Medicine, 32*(12), 1315-1322. https://doi.org/10.1007/s11606-017-4150-7

Chaumont, M., van de Borne, P., Bernard, A., Van Muylem, A., Deprez, G., Ullmo, J., Starczewska, E., Briki, R., de Hemptinne, Q., Zaher, W., & Debbas, N. (2019). Fourth generation e-cigarette vaping induces transient lung inflammation and gas exchange disturbances: Results from two randomized clinical trials. *American Journal of Physiology - Lung Cellular and Molecular Physiology, 316*(5), L705-L719. https://doi.org/10.1152/ajplung.00492.2018

Coppeta, L., Magrini, A., Pietroiusti, A., Perrone, S., & Grana, M. (2018). Effects of smoking electronic cigarettes on pulmonary function and environmental parameters. *Open Public Health Journal, 11*(1), 360-368. https://doi.org/10.2174/1874944501811010360

Cravo, A. S., Bush, J., Sharma, G., Savioz, R., Martin, C., Craige, S., & Walele, T. (2016, Nov 15). A randomised, parallel group study to evaluate the safety profile of an electronic vapour product over 12 weeks. *Regul Toxicol Pharmacol, 81 Suppl 1*, S1-s14. https://doi.org/10.1016/j.yrtph.2016.10.003

D'Ruiz, C. D., O'Connell, G., Graff, D. W., & Yan, X. S. (2017). Measurement of cardiovascular and pulmonary function endpoints and other physiological effects following partial or complete substitution of cigarettes with electronic cigarettes in adult smokers. *Regulatory Toxicology and Pharmacology, 87*, 36-53. https://doi.org/10.1016/j.yrtph.2017.05.002

Flouris, A. D., Chorti, M. S., Poulianiti, K. P., Jamurtas, A. Z., Kostikas, K., Tzatzarakis, M. N., Wallace Hayes, A., Tsatsaki, A. M., & Koutedakis, Y. (2013). Acute impact of active and passive electronic cigarette smoking on serum cotinine and lung function. *Inhalation Toxicology, 25*(2), 91-101. https://doi.org/10.3109/08958378.2012.758197

Kotoulas, S. C., Pataka, A., Domvri, K., Spyratos, D., Katsaounou, P., Porpodis, K., Fouka, E., Markopoulou, A., Passa-Fekete, K., Grigoriou, I., Kontakiotis, T., Argyropoulou, P., & Papakosta, D. (2020). Acute effects of e-cigarette vaping on pulmonary function and airway inflammation in healthy individuals and in patients with asthma. *Respirology, 25*(10), 1037-1045. https://doi.org/10.1111/resp.13806

Lappas, A. S., Tzortzi, A. S., Konstantinidi, E. M., Teloniatis, S. I., Tzavara, C. K., Gennimata, S. A., Koulouris, N. G., & Behrakis, P. K. (2018). Short-term respiratory effects of e-cigarettes in healthy individuals and smokers with asthma. *Respirology*, *23*(3), 291-297. https://doi.org/10.1111/resp.13180

Palamidas, A., Tsikrika, S., Katsaounou, P. A., Vakali, S., Gennimata, S. A., Kaltsakas, G., Gratziou, C., & Koulouris, N. (2017). Acute effects of short term use of ecigarettes on Airways Physiology and Respiratory Symptoms in Smokers with and without Airway Obstructive Diseases and in Healthy non smokers. *Tob Prev Cessat, 3,* 5. https://doi.org/10.18332/tpc/67799

Polosa, R., Morjaria, J. B., Caponnetto, P., Caruso, M., Campagna, D., Amaradio, M. D., Ciampi, G., Russo, C., & Fisichella, A. (2016, Feb). Persisting long term benefits of smoking abstinence and reduction in asthmatic smokers who have switched to electronic cigarettes. *Discov Med*, *21*(114), 99-108.

Polosa, R., Morjaria, J. B., Caponnetto, P., Prosperini, U., Russo, C., Pennisi, A., & Bruno, C. M. (2016, Dec 16). Evidence for harm reduction in COPD smokers who switch to electronic cigarettes. *Respir Res, 17*(1), 166. https://doi.org/10.1186/s12931-016-0481-x

Polosa, R., Morjaria, J. B., Prosperini, U., Busà, B., Pennisi, A., Malerba, M., Maglia, M., & Caponnetto, P. (2020). COPD smokers who switched to e-cigarettes: health outcomes at 5-year follow up. *Therapeutic Advances in Chronic Disease, 11*. https://doi.org/10.1177/2040622320961617

Polosa, R., Morjaria, J. B., Prosperini, U., Russo, C., Pennisi, A., Puleo, R., Caruso, M., & Caponnetto, P. (2018). Health effects in COPD smokers who switch to electronic cigarettes: a retrospective-prospective 3-year follow-up. *Int J Chron Obstruct Pulmon Dis, 13*, 2533-2542. https://doi.org/10.2147/COPD.S161138

Pulvers, K., Nollen, N. L., Rice, M., Schmid, C. H., Qu, K., Benowitz, N. L., & Ahluwalia, J. S. (2020). Effect of Pod e-Cigarettes vs Cigarettes on Carcinogen Exposure Among African American and Latinx Smokers: A Randomized Clinical Trial. *JAMA network open, 3*(11), e2026324. https://doi.org/10.1001/jamanetworkopen.2020.26324

Suhling, H., Welte, T., & Fuehner, T. (2020). Three patients with acute pulmonary damage following the use of E-cigarettes—A case series. *Deutsches Arzteblatt International, 117*(11), 177-182. https://doi.org/10.3238/arztebl.2020.0177

Vardavas, C. I., Anagnostopoulos, N., Kougias, M., Evangelopoulou, V., Connolly, G. N., & Behrakis, P. K. (2012). Short-term pulmonary effects of using an electronic cigarette: Impact on respiratory flow resistance, impedance, and exhaled nitric oxide. *Chest, 141*(6), 1400-1406. https://doi.org/10.1378/chest.11-2443

Veldheer, S., Yingst, J., Midya, V., Hummer, B., Lester, C., Krebs, N., Hrabovsky, S., Wilhelm,

A., Liao, J., Yen, M. S., Cobb, C., Eissenberg, T., & Foulds, J. (2019, Apr). Pulmonary and other health effects of electronic cigarette use among adult smokers participating in a randomized controlled smoking reduction trial. *Addict Behav, 91,* 95-101. https://doi.org/10.1016/j.addbeh.2018.10.041

Walele, T., Bush, J., Koch, A., Savioz, R., Martin, C., & O'Connell, G. (2018, Feb). Evaluation of the safety profile of an electronic vapour product used for two years by smokers in a real-life setting. *Regul Toxicol Pharmacol, 92*, 226-238. https://doi.org/10.1016/j.yrtph.2017.12.010

### I. Data extraction form

#### Data Extraction Form March 1

Reviewer: Date: Verified on:

Title:

Year:

Journal: DOI: PMID:

1st Author:

Affiliations:

COI Declaration:

2nd Author

Affiliations:

COI Declarations:

[all additional authors]

Corresponding author and email:

Additional/supplementary files [identify or NONE]

Published protocol or trial registry [identify or NONE]

Funder:

Research question (or goal if no stated question)

Study design:

Location (Country):

Setting:

Participants:

- Age
- Sex
- Exclusion criteria
- Smoking status/history/current use
- Other tobacco products reported
- ENDS history
- Disease status
- Other descriptors (economic status, etc.)

Recruitment and Compensation

Randomization/assignment process

Statistical power

Intervention

- ENDS device and liquid
- ENDS training
- Frequency of use
- Duration of exposure
- Concurrent use of tobacco
- Follow-up periods
- Drop-outs
- Fidelity including relapse to cigarette use during study

Tests/data

- Data collection methods

Named tests

Test not eligible for data extraction

Testing protocols including pretest abstinence period

Observational data (disease symptoms)

Self-report data

Verification of tobacco abstinence

Tests results/data [for each follow-up period]

Tests

Observational data

Self-report data

Dual user and exclusive users reported separately or combined

Secondary outcomes (tobacco cessation, ENDS cessation, tobacco relapse)

Statistical tests

Statistical analyses

Limitations identified by authors

Recommendations for future research

Conclusions(s) quotations (with page number)

Reviewer Comments:

### J. Data discrepancies form

Reviewer: Date: Verified on:

Author (year):

Indicate status:

No discrepancies observed.

Discrepancies in data between the abstract and the study text:

Discrepancies within the text (compare all references in the text to the data):

Discrepancies between the text and figure:

Discrepancies between the text and table:

Discrepancies in number of participants:

Study corresponding author contacted on date and text of email:

(If no reply): Other authors and cc of journal editor on date and text of email:

Review paper: flagged in study table, noted in QA.

K. Protocol discrepancies form

Reviewer  Date  Verified on

Author (year)

Status (select):

No protocol/trial registry reported in study.

Protocol/trial registry reported in study, not published.

Protocol/trial registry published.

Deviations from protocol/trial registry reported by study authors:

Deviations from published protocol/trial registry observed by reviewer:

L. Bias report

Reviewer   date   verification

Author (year)

Reporting Biases

Spin Bias

Data-dredging bias

Hypothetical bias

All’s well literature bias

Ascertainment bias

Biases of rhetoric

Compliance bias

Confirmation bias

Detection bias

Hot stuff bias

Industry Sponsorship Bias

Misclassification bias

One-sided reference bias

Partial reference bias

Performance bias

Popularity bias

Prevalence-incidence (Neyman) bias

Selection bias

Substitution game bias

Volunteer bias

Wrong sample size bias

### PRISMA-P 2015 Checklist

This checklist has been adapted for use with protocol submissions to *Systematic Reviews* from Table 3 in Moher D et al: Preferred reporting items for systematic review and meta-analysis protocols (PRISMA-P) 2015 statement. *Systematic Reviews* 2015 4:1

| Section/topic | # | Checklist item | Information reported | | Page number |
|---|---|---|---|---|---|
| | | | Yes | No | |
| **ADMINISTRATIVE INFORMATION** | | | | | |
| **Title** | | | | | |
| Identification | 1a | Identify the report as a protocol of a systematic review | X | | 1 |
| Update | 1b | If the protocol is for an update of a previous systematic review, identify as such | | | Not applicable |
| **Registration** | 2 | If registered, provide the name of the registry (e.g., PROSPERO) and registration number in the Abstract | X | | Submitted 9 |
| **Authors** | | | | | |
| Contact | 3a | Provide name, institutional affiliation, and e-mail address of all protocol authors; provide physical mailing address of corresponding author | X | | 1 |
| Contributions | 3b | Describe contributions of protocol authors and identify the guarantor of the review | X | | 21 |
| **Amendments** | 4 | If the protocol represents an amendment of a previously completed or published protocol, identify as such and list changes; otherwise, state plan for documenting important protocol amendments | X | | 9 |
| **Support** | | | | | |
| Sources | 5a | Indicate sources of financial or other support for the review | X | | 21 |
| Sponsor | 5b | Provide name for the review funder and/or sponsor | X | | 21 |
| Role of sponsor/funder | 5c | Describe roles of funder(s), sponsor(s), and/or institution(s), if any, in developing the protocol | X | | 21 |
| **INTRODUCTION** | | | | | |
| **Rationale** | 6 | Describe the rationale for the review in the context of what is already known | X | | 7 |
| **Objectives** | 7 | Provide an explicit statement of the question(s) the review will address with reference to participants, interventions, comparators, and outcomes (PICO) | X | | 7-8 |
| **METHODS** | | | | | |

| Section/topic | # | Checklist item | Information reported Yes | Information reported No | Page number |
|---|---|---|---|---|---|
| **Eligibility criteria** | 8 | Specify the study characteristics (e.g., PICO, study design, setting, time frame) and report characteristics (e.g., years considered, language, publication status) to be used as criteria for eligibility for the review | X | | 7-8, 11-12 |
| **Information sources** | 9 | Describe all intended information sources (e.g., electronic databases, contact with study authors, trial registers, or other grey literature sources) with planned dates of coverage | X | | 10-11, 13 |
| **Search strategy** | 10 | Present draft of search strategy to be used for at least one electronic database, including planned limits, such that it could be repeated | X | | Suppl. Materials |
| ***STUDY RECORDS*** | | | | | |
| Data management | 11a | Describe the mechanism(s) that will be used to manage records and data throughout the review | X | | 11 |
| Selection process | 11b | State the process that will be used for selecting studies (e.g., two independent reviewers) through each phase of the review (i.e., screening, eligibility, and inclusion in meta-analysis) | X | | 12 |
| Data collection process | 11c | Describe planned method of extracting data from reports (e.g., piloting forms, done independently, in duplicate), any processes for obtaining and confirming data from investigators | X | | 13, Suppl. Materials |
| **Data items** | 12 | List and define all variables for which data will be sought (e.g., PICO items, funding sources), any pre-planned data assumptions and simplifications | X | | 7-8, 13, Suppl. Materials |
| **Outcomes and prioritization** | 13 | List and define all outcomes for which data will be sought, including prioritization of main and additional outcomes, with rationale | X | | 7-8, 13, Suppl. Materials |
| **Risk of bias in individual studies** | 14 | Describe anticipated methods for assessing risk of bias of individual studies, including whether this will be done at the outcome or study level, or both; state how this information will be used in data synthesis | X | | 14-15, Suppl. Materials |
| ***DATA*** | | | | | |
| **Synthesis** | 15a | Describe criteria under which study data will be quantitatively synthesized | X | | 15 |
| | 15b | If data are appropriate for quantitative synthesis, describe planned summary measures, methods of handling data, and methods of combining data from studies, | | | Not applicable |

| Section/topic | # | Checklist item | Information reported | | Page number |
|---|---|---|---|---|---|
| | | | Yes | No | |
| | | including any planned exploration of consistency (e.g., $I^2$, Kendall's tau) | | | |
| | 15c | Describe any proposed additional analyses (e.g., sensitivity or subgroup analyses, meta-regression) | X | | 15 |
| | 15d | If quantitative synthesis is not appropriate, describe the type of summary planned | X | | 15 |
| **Meta-bias(es)** | 16 | Specify any planned assessment of meta-bias(es) (e.g., publication bias across studies, selective reporting within studies) | X | | 14, 15 |
| **Confidence in cumulative evidence** | 17 | Describe how the strength of the body of evidence will be assessed (e.g., GRADE) | X | | 15 |
